# A territory-wide study of early COVID-19 outbreak in Hong Kong community: A clinical, epidemiological and phylogenomic investigation

**DOI:** 10.1101/2020.03.30.20045740

**Authors:** Kenneth Siu-Sing Leung, Timothy Ting-Leung Ng, Alan Ka-Lun Wu, Miranda Chong-Yee Yau, Hiu-Yin Lao, Ming-Pan Choi, Kingsley King-Gee Tam, Lam-Kwong Lee, Barry Kin-Chung Wong, Alex Yat-Man Ho, Kam-Tong Yip, Kwok-Cheung Lung, Raymond Wai-To Liu, Eugene Yuk-Keung Tso, Wai-Shing Leung, Man-Chun Chan, Yuk-Yung Ng, Kit-Man Sin, Kitty Sau-Chun Fung, Sandy Ka-Yee Chau, Wing-Kin To, Tak-Lun Que, David Ho-Keung Shum, Shea Ping Yip, Wing Cheong Yam, Gilman Kit-Hang Siu

## Abstract

Initial cases of COVID-19 reported in Hong Kong were mostly imported from China. However, most cases reported in February 2020 were locally-acquired infections, indicating local community transmissions. We extracted the demographic, clinical and epidemiological data from 50 COVID-19 patients, who accounted for 53.8% of the cases in Hong Kong by February 2020. Whole-genome sequencing of the SARS-CoV-2 were conducted to determine the phylogenetic relatedness and transmission dynamics. Only three (6.0%) patients required ICU admission. Phylogenetic analysis identified six transmission clusters. All locally-acquired cases harboured a common mutation *Orf3a* G251V and were clustered in two subclades in global phylogeny of SARS-CoV-2. The estimated time to the most recent common ancestor of local COVID-2019 outbreak was December 24, 2019 with an evolutionary rate of 3.04×10^−3^ substitutions per site per year. The reproduction number value was 1.84. Social distancing and vigilant epidemiological control are crucial to the containment of COVID-19 transmission.

**Article summary lines:** A combined epidemiological and phylogenetic analysis of early COVID-19 outbreak in Hong Kong revealed that a SARS-CoV-2 variant with *ORF3a* G251V mutation accounted for all locally acquired cases, and that asymptomatic carriers could be a huge public health risk for COVID-19 control.

## Introduction

“Coronavirus disease 2019(COVID-19)” refers to a cluster of viral pneumonia cases initially from Wuhan, Hubei Province, China since December 2019. The aetiology was unknown during the early stage of the outbreak until Chinese scientists isolated a novel coronavirus, SARS-CoV-2, on January 7, 2020 and performed genome sequencing(1).

Fever was the main symptom of COVID-19 with about one-third of the patients presenting acute respiratory distress syndrome(ARDS). About 16% of the patients were in severe condition on admission, and the estimated mortality rate was 1.4%(2). Sustained human-to-human transmission was confirmed upon identification of cases clustering among families and transmission from patients to healthcare workers(3, 4), which triggered China’s urgent public health actions and international concern.

As of February 28, 2020, 78,824 patients have been diagnosed to have COVID-19 in Mainland China with 2,788 deaths. The disease also spread to 50 other countries (5). The World Health Organization(WHO) declared COVID-19 pandemic in March 2020. The first imported case in Hong Kong was on January 23, 2020, who was a Mainland China resident travelled to Hong Kong from Wuhan via Shenzhen by High-Speed Rail. The first local case with unknown source, i.e. patient who had no travel record during the 14-day incubation period, was reported on February 4, 2020(6).

By February 28, 2020, there were 93 COVID-19 cases in Hong Kong with at least 70(75·3%) cases being local cases and their close contacts(6, 7). Secondary and tertiary transmissions were observed in some case clusters. As most index cases of these clusters have unknown sources, hidden transmission chain was believed to have established in the community.

Here, we report the demographic, clinical and epidemiological data of 50 hospitalised patients, who accounted for 53.8% of COVID-19 cases in Hong Kong at the data cut-off point (February 28, 2020), including three imported cases and six transmission clusters of local infections. With the combined use of Nanopore sequencing and Illumina sequencing, viral genomes of all these cases were characterised. Phylogenetic and molecular evolutionary analyses were performed to determine the transmission linkage and the evolutionary rate of COVID-19 cases in the community.

## Methods

### Cases

For this retrospective, multi-centre study, we intentionally enrolled the positive cases of COVID-19 that were laboratory-confirmed at four public hospital clusters managed under the Hospital Authority of Hong Kong, namely Hong Kong East Cluster, Kowloon East Cluster, Kowloon West Cluster, and New Territories West Cluster, from January 26 to February 28, 2020.

Sputum specimens and throat swabs pooled with nasopharyngeal aspirates were collected from patients who fulfilled the reporting or enhanced surveillance criteria on admission(8). Laboratory-confirmed infection was defined when SARS-CoV-2 was detected by real-time reverse transcription polymerase chain reactions(RT-PCRs), which amplified the envelope(*E*) gene and RNA-dependent RNA polymerase(*RdRp*) gene(9).

Demographic, clinical, and microbiological data were obtained from patients’ medical records. Epidemiological information was retrieved from the Centre for Health Protection(CHP) of the Department of Health(6) and the website wars.vote4.hk - Coronavirus in HK(7). The definitions of clinical symptoms and complications are based on the WHO guidance(10). We adopted the case numbering system of CHP, which was based on the date of case confirmation.

This study was approved by the Institutional Review Boards of The Hong Kong Polytechnic University(RSA20021) and the public hospitals involved(HKECREC-20200014;KCC/KEC-20200070;KWC-20200040;NTWC-20200038).

### Specimen preparation

The respiratory specimens were centrifuged at 16,000*g* for 2 minutes. Total nucleic acid was extracted from supernatant using MagNA Pure 96 System(Roche Diagnostics, Germany) or NucliSENS^®^ easyMAG^®^(bioMérieux, The Netherlands) according to manufacturers’ instructions. DNase treatment was done by TURBO DNA-free Kit(ThermoFisher Scientific, USA) to remove residual host DNA.

### Reverse Transcription and viral genome amplification using multiplex PCR

DNase-treated RNA was reverse-transcribed using random hexamers and SuperScript IV reverse transcriptase(Invitrogen, USA) as previously described(11). Viral cDNA was then amplified using two PCRs containing tiled, multiplexed primers(table S1) described in the ARTIC network(12). Details of the multiplex PCR are provided in the Supplemental Material.

### Nanopore MinION Sequencing

Ligation-based 1D sequencing(SQK-LSK109, ONT, UK) was carried out according to manufacturer’s instructions. Briefly, multiplex PCR amplicons of each sample were normalised to 1 ng/µL prior to end-repairing and native barcode ligation(EXP-NBD104/114, ONT, UK). Barcoded samples were pooled and ligated to AMII sequencing adaptor. Sequencing was performed with Nanopore MinION device using R9.4.1 flow cell for 48 hours.

### Illumina MiSeq sequencing

Multiplex PCR amplicons were subjected to library preparation and dual-indexing using KAPA HyperPrep Kit and Unique Dual-Indexed Adapter Kit(Roche Applied Science, US) according to manufacturer’s instructions. Ligated libraries were enriched by 6-cycle PCR amplification followed by purification and size selection using AMPure XP beads(Beckman Coulter, USA). The pooled library was sequenced with MiSeq Reagent Kit V2 Nano on Illumina MiSeq System.

### Bioinformatic analysis

Nanopore sequencing data were analysed using modified Artic Network nCoV-2019 novel coronavirus bioinformatics protocol(Supplemental Material)(13).

Illumina sequencing reads were mapped with reference to respective draft genome of each sample constructed from Nanopore data. Variants were called using freebayes(v1.0.0) with haploid decoding and minimum base quality set at Q30. Consensus genomes were constructed by GATK(v4.1.4.1) based on the VCF file(14). SPAdes genome assembler(v3.14.0) and minimap2(v2.17) were used to combine Nanopore and Illumina sequencing results for *de novo* assembly and identify the sequence of the unmapped gap regions. The sequences have been submitted to the GenBank with accession no. MT232662-MT232711.

### Genomic and Phylogenetic analysis

To identify the amino acid change caused by each single-nucleotide polymorphism(SNP), consensus genome of each specimen was BLASTed against the reference NC_045512.2 using BlastX. Non-synonymous mutations were identified using custom Python script.

Phylogenetic tree was constructed based on the consensus genomes with PhyML(v3.0) using maximum likelihood algorithm. Best-fitting substitution model was selected by Akaike information criteria, in which general time reversible model with fixed proportion of invariable sites(+I) was selected(15). Bootstrap replicates was set at 1000×, and maximum-likelihood phylogenetic tree was rooted on the earliest published genome (accession no.: NC_045512.2). Transmission cluster was defined by clear epidemiological and onset-time relationship. Meanwhile, an additional 273 SARS-CoV-2 genomes were downloaded from GISAID Severe acute respiratory syndrome coronavirus 2 data hub(16). Together with the genomes from this study, phylogenetic tree was constructed using fast likelihood-based aLRT SH-like method and rooted on SARS-CoV-2 genome NC_045512.2.

### Estimation of evolutionary rate and divergence time of transmission

To reconstruct the evolutionary model of COVID-19 cases in Hong Kong, Bayesian interference through Markov Chain Monte Carlo(MCMC) framework was implemented in BEAST(v2.6.2)(17). Bayesian phylodynamic analysis was performed using both strict clock and relaxed clock model with coalescent exponential growth tree priors. MCMC chains were run for 1 × 10^9^ generations and sampled every 500 steps. Bayesian output was analysed after the results were visualised by Tracer(v1.7.1)(18). All parameters had an effective sample size of >200, indicating sufficient sampling.

## Results

Fifty COVID-19 patients were included with 54·0% being female and the mean age was 55·2 years (table 1). Three were considered as imported cases as the patients stayed in Wuhan before travelling to Hong Kong in mid-January. Four travelled to other regions where COVID-19 active community transmission was not confirmed by that time. These cases were considered as possibly local infection. The other 43 were defined as local infection with no recent travel history. Eighteen(36·0%) had chronic illnesses, of which cardiovascular and cerebrovascular diseases were the most common conditions(table 1). In total, 74·0% of the patients presented with cough. Fever was presented in 58·0% of the patients on admission, but gradually developed in 64·0% patients during hospitalisation. Other less common symptoms include muscle-ache(25·0%), sore throat(24·0%), shortness of breath(24·0%), and diarrhoea(14·3%)(table 2). Two(4·0%) patients were asymptomatic throughout the study period. Regarding radiological examination, 27(54·0%) showed bilateral pneumonia, 11(22·0%) unilateral pneumonia, and 17(34·7%) multiple mottling and ground-glass opacity. None of the patients was co-infected with other respiratory viruses or fungi. Two patients were culture-positive for *Klebsiella aerogenes* and *Ralstonia pickettii* in their sputum specimens. Both presented with ARDS and acute respiratory injury accompanied with septic shock or acute renal injury, and required ICU admission.

**Table 1:**
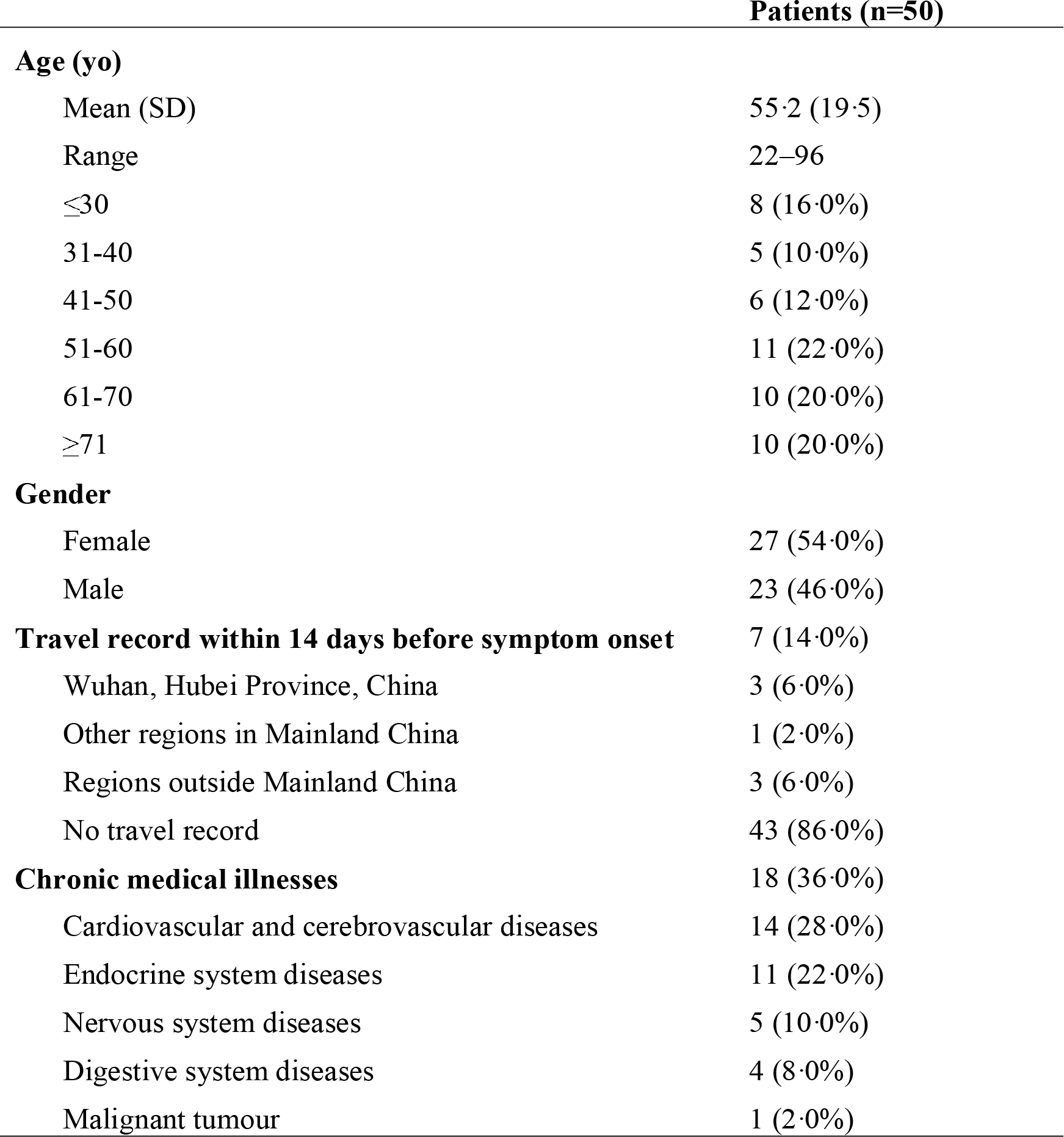
Demographics, travel record and baseline medical history of 50 patients recruited in this study.

**Table 2:**
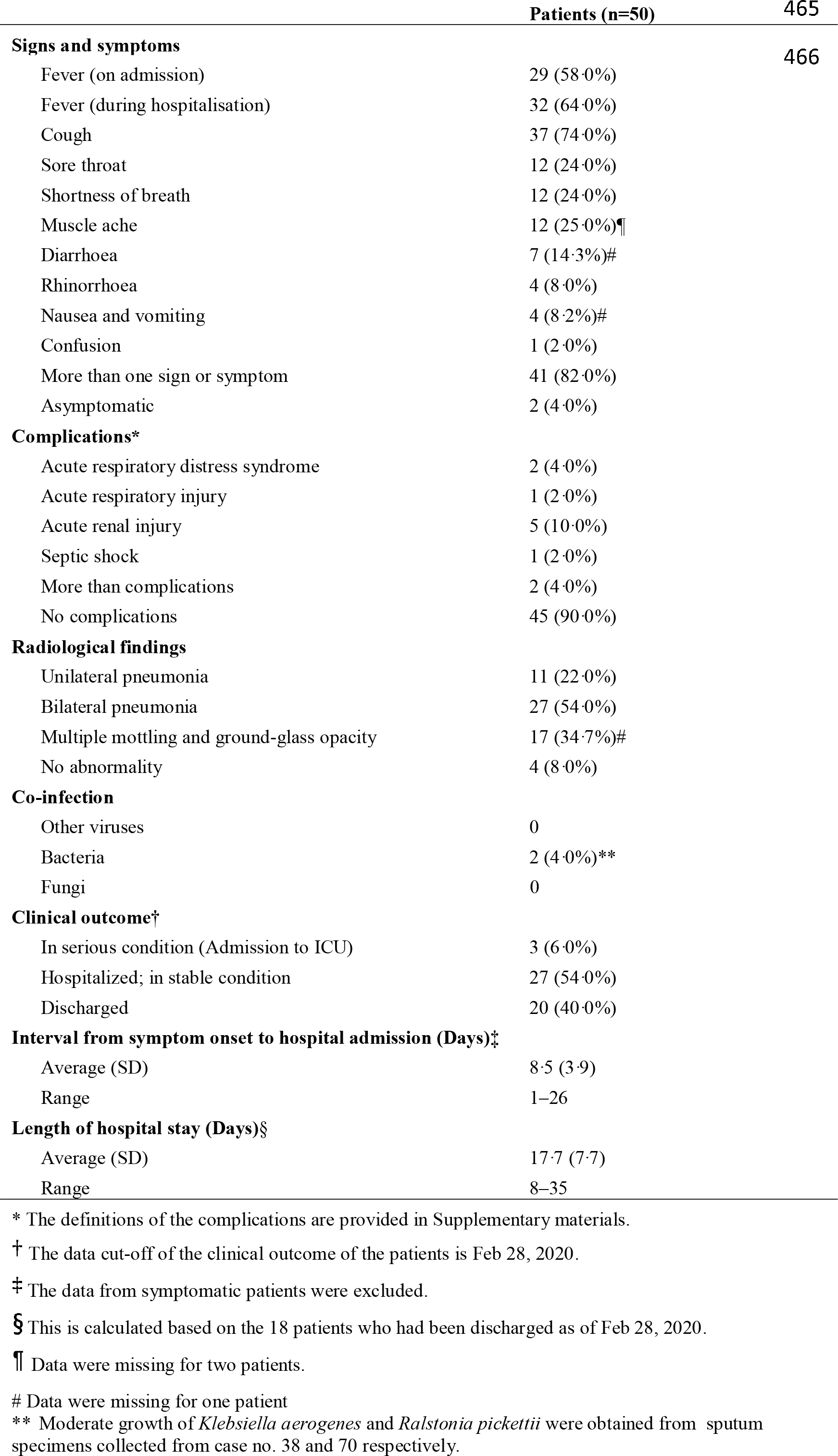
Clinical characteristics and outcomes of 50 patients recruited in this study.

Of the 50 cases, 42(84·0%) could be clustered based on their epidemiological linkages(figure 1). Six transmission clusters(Clusters 1–6) were identified. Cluster 1 involved a family of four members. The father, who travelled to Guangdong, China in late January 2020, was believed to infect his wife and subsequently their daughter and son-in-law in a family gathering. Clusters 2 and 3 were family clusters of local infection with unknown source. Both clusters involved three household members without recent travel history. Cluster 4 was a super-spreading event(SSE) associated with a barbecue and hotpot party of 19 family members in late January. Ten had developed symptoms two days after the party. A colleague of one infected member, who was not present at the party, was also diagnosed to have COVID-19. Cluster 5 initiated from a resident of a public estate, who was diagnosed on January 30. Eleven days later, three members of a household living in the same building as, but 10-storey below from, the index case were also infected. Two household members attended a family gathering of 29 people at a Chinese seafood restaurant during their incubation period. Three were diagnosed consecutively around two weeks after the gathering. Additionally, a Filipino domestic helper of one infected member, who was absent in the family gathering, was also infected. The earliest reported case of Cluster 6 was a woman who visited a Buddhist worship hall during Chinese New Year holidays. Later, eight individuals who visited the same Buddhist worship hall during this period were diagnosed. At the data cut-off point, at least four other household members who had never been to the worship hall were also tested positive for SARS-CoV-2.

**Figure 1.**
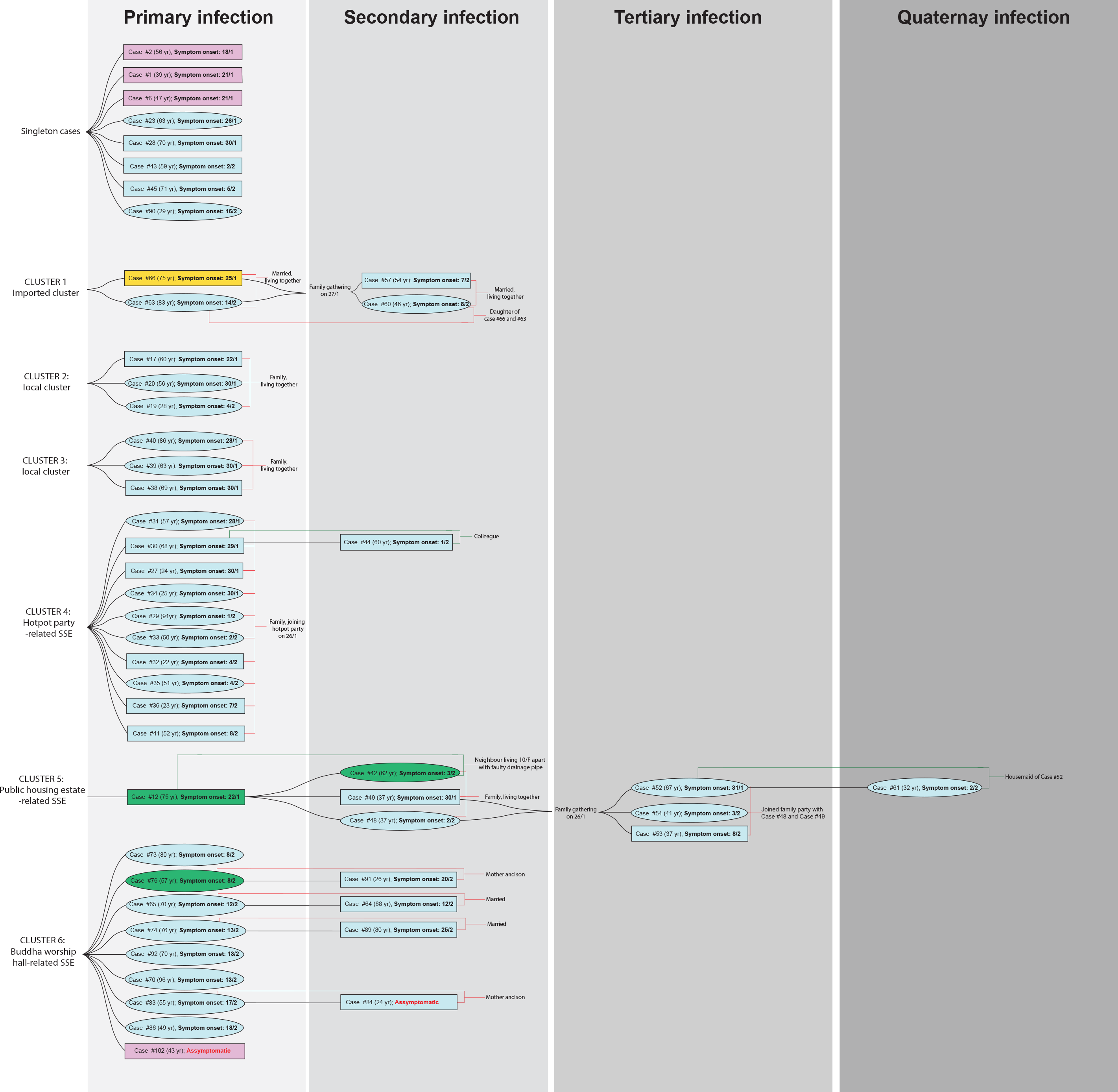
Demographics of patients included in this study. A rectangular box stands for male and eclipse for female. Travel history within the 14-day incubation period prior to symptom onset is highlighted in 1) Cyan for local case without travel history; 2) Pink for travel history to Wuhan within 14-days from symptom onset; 3) Orange for travel history to other regions in Mainland China, and 4) Green for travel history to regions outside Mainland China. The case numbers are those used by the Centre of Health Protection, Department of Health, Hong Kong.^6^ Within each cluster, the cases of primary infection (leftmost column) are arranged from top to bottom in the order of the date (dd/m) of symptom onset.

Consensus genomes of all 50 cases were constructed based on Nanopore sequencing and refined by Illumina sequencing. On average, 62,387 and 18,747 reads were obtained per genome with 550× and 132× coverage for Nanopore and Illumina platforms respectively. The consensus genome size was ∼29.9 kbp with GC content ∼38%. The genomes were highly conserved with the first SARS-CoV-2 genome with an average sequence identity of 99·98%(range: 99·94%–100·0%). A total of 64 nonsynonymous substitutions were identified from all 50 genomes(table 3). G251V in *Orf3a* is the most frequent amino acid substitution with 44/50(88·0%) of the samples harbouring this mutation, followed by *Orf1ab* H3233Y(30/50; 60·0%) and *S* gene L8V(27/50; 54·0%).

**Table 3:**
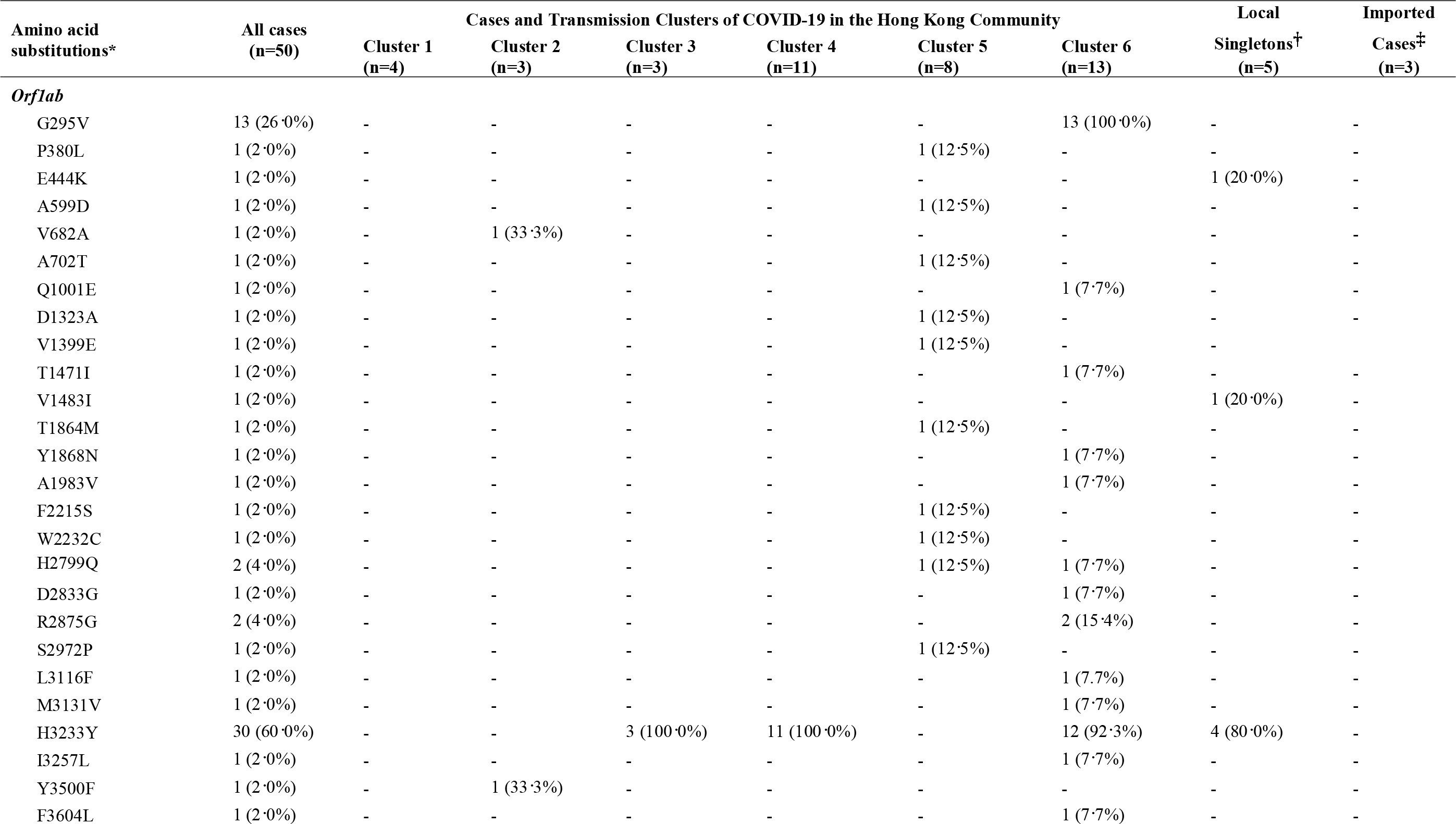
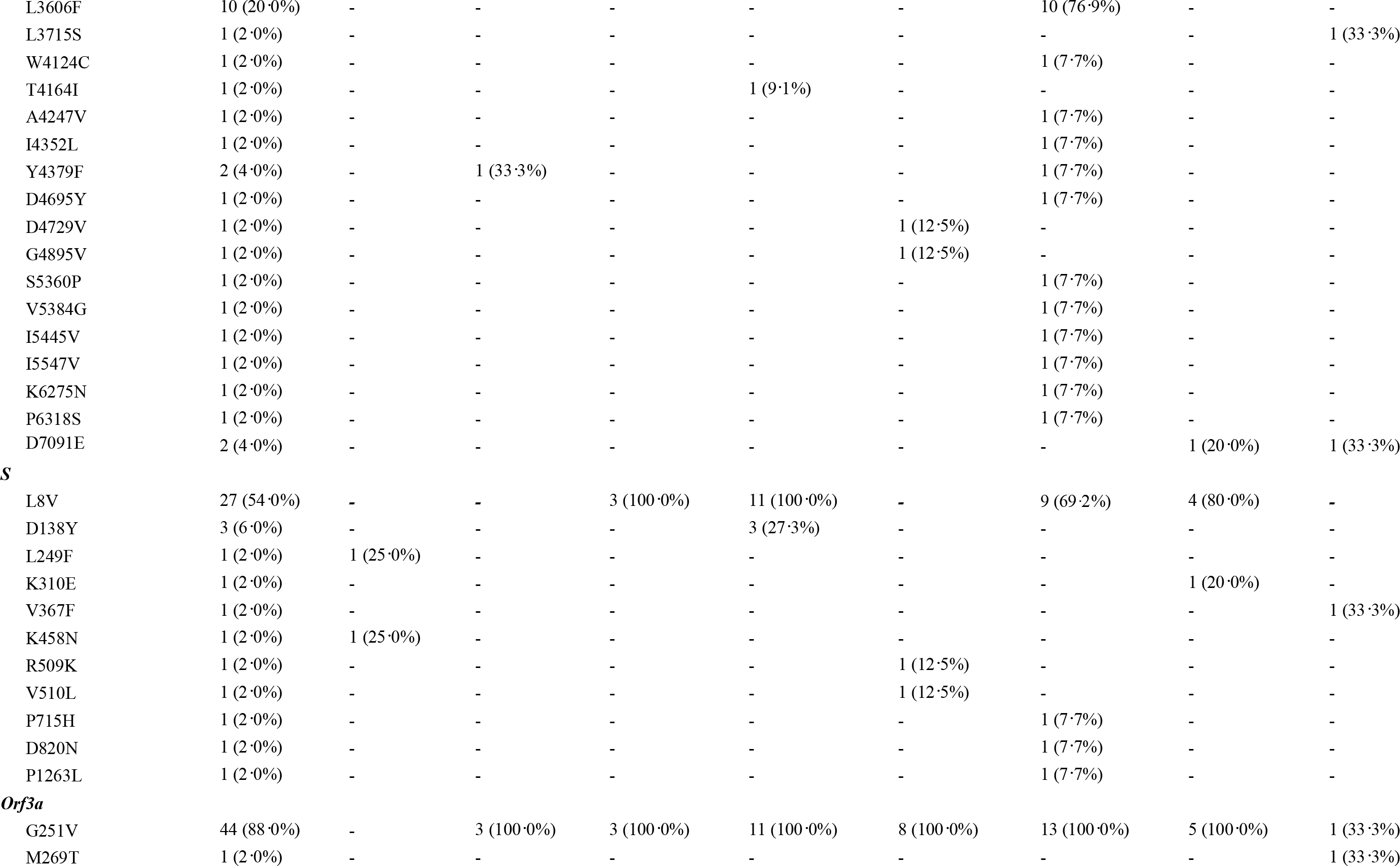
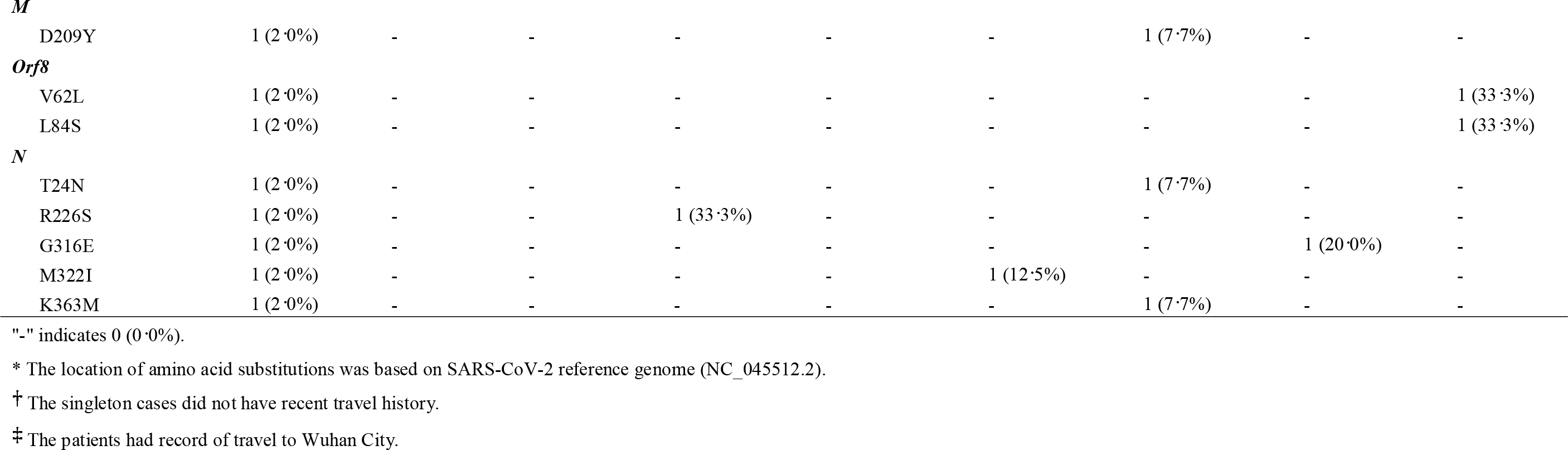
Amino acid substitutions identified in SARS-CoV-2 isolated from the 50 patients in this study.

Genomic-wide SNPs were used to contextualise phylogenetic placement of Hong Kong strains in SARS-CoV-2global phylogeny. Compared to SARS-CoV-2 strains isolated from other regions, Hong Kong strains showed limited genomic variability, and tended to aggregate in two clusters(figure 2). In the first cluster, a total of 13 Hong Kong strains formed a subclade(yellow box) with the strains from France(n=5), Singapore(n=4), Australia(n=2), South Korea(n=2), Sweden(n=1) and USA(n=1). The second cluster composed of 31 Hong Kong strains, which formed another distinct subclade(pink box) with the strains from Japan(n=3), China(n=2), England(n=2), Australia(n=2) and New Zealand(n=1).

**Figure 2.**
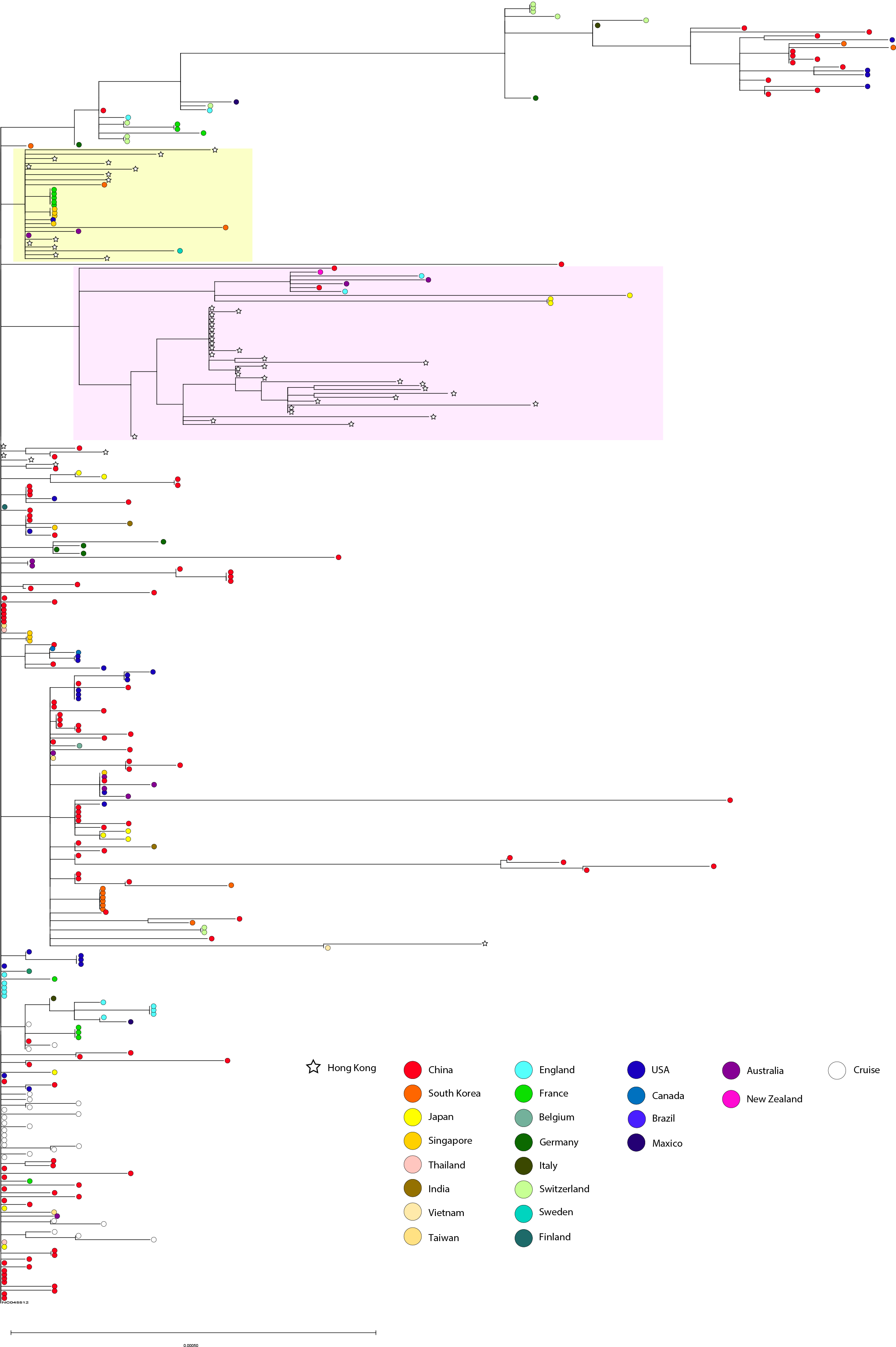
Maximum-likelihood phylogenetic tree constructed based on 50 SARS-CoV-2 genomes in this study and genomes collected from GISAID data hub. A total of 273 worldwide genomes as of February 28, 2020 were randomly selected and downloaded from GISAID Global Cases COVID-19 database. The phylogenetic tree was built with the samples collected in this study based on fast likelihood-based aLRT SH-like method and rooted on the earliest published genome of SARS-CoV-2 (accession no.: NC_045512.2). The Hong Kong strains showed limited genetic variability and tended to aggregate in two subclades, which were highlighted in yellow box and pink box respectively.

Regarding the phylogenetic relationship among the Hong Kong cases, clustering of samples was consistent with the epidemiological linkage(figure 3). Four distinctive nodes were identified. The first node belonged to two imported cases and the cases in Cluster 1 of which the genomes had very close genetic distance(0–2 SNPs) with the reference genome. The second node represented cases in Cluster 5, of which four patients lived in different apartments of the same building in a public housing estate, and were believed to have transmitted the virus to other members in a subsequent family gathering. The third node was associated with the cases in Cluster 4 which believed to originate from a family hotpot gathering. Notably, the genomes of Cluster 4 were highly similar to those in Cluster 3 and two singleton cases(i.e. Case 23 and Case 43), which shared the same missense mutations at L8V in the *S* gene, H3233Y in *Orf1ab* and G251V in *Orf3a*. Finally, the fourth node belonged to the SSE occurring in Buddhist worship hall(Cluster 6), in which *Orf1ab* G295V and *Orf3a* G251V were identified in all the strains in the cluster.

**Figure 3.**
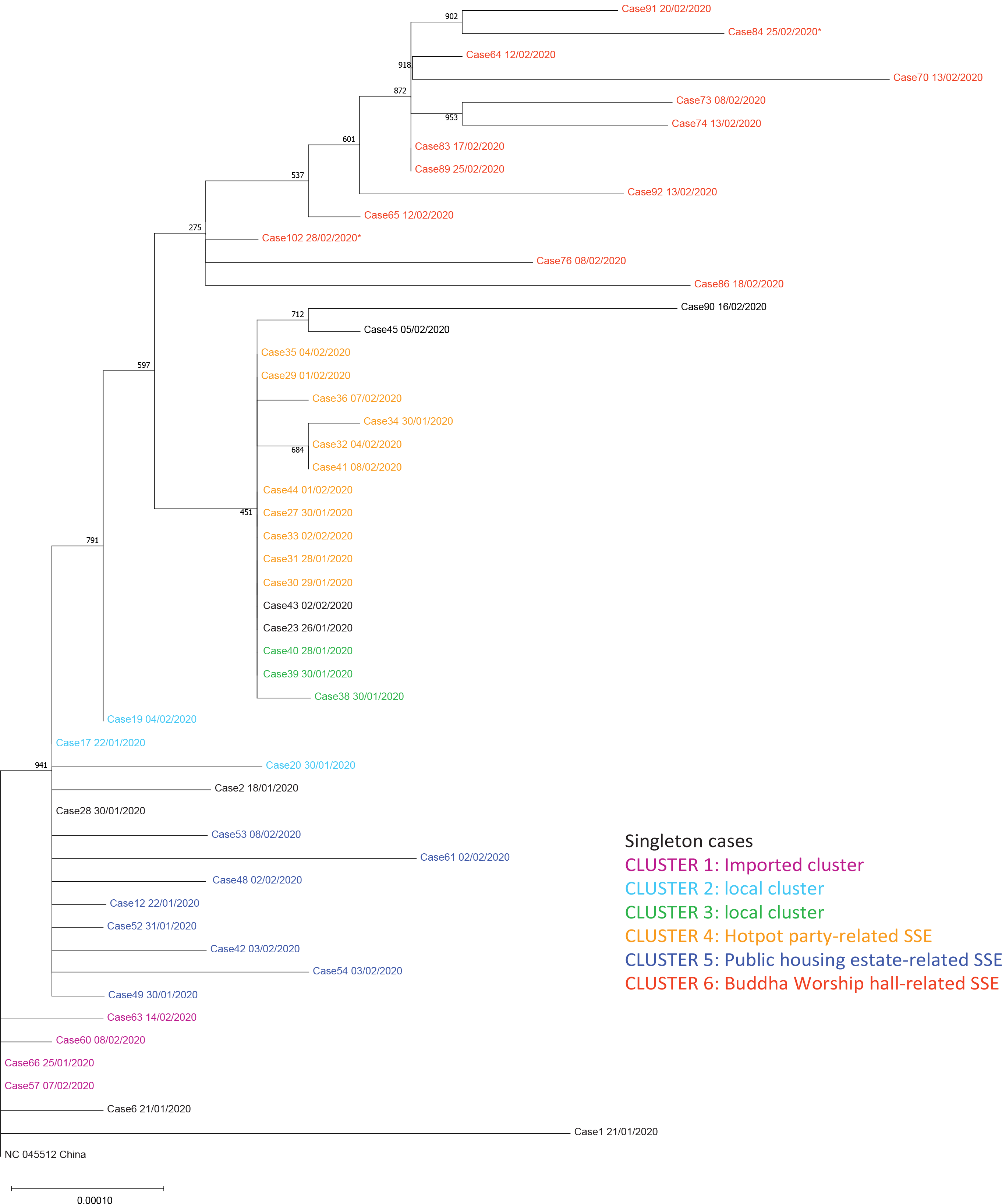
Maximum-likelihood phylogenetic tree of the 50 COVID-19 cases in this study. The tree was constructed using all 50 COVID-19 cases included in this study and rooted on the earliest published genome of SARS-CoV-2 (accession no.: NC_045512.2). Bootstrap value was set at 1000×. Samples were colour-coded by epidemiological link as follows: 1) Magenta represents Cluster 1: Imported cluster; 2) Cyan represents Cluster 2: unknown source; 3) Green represents Cluster 3: local; 4) Orange represents Cluster 4: Hotpot party-related Superspreading event (SSE); 5) Blue represents Cluster 5: Public housing estate-related SSE; and 6) Red represents Cluster 6: Buddha worship hall-related SSE. Case 84 and Case 102 were asymptomatic at the time of sample collection, and were marked with asterisks (*) in the diagram. Each case is indicated with a case number (see figure 1) followed by the date (dd/mm/yyyy) of symptom onset.

According to Bayesian time-scaled phylodynamic analysis, estimated tMRCA of COVID-2019 outbreak in Hong Kong was December 24, 2019(95% Bayesian credible interval [BCI]: December 11, 2019 to January 5, 2020) with an evolutionary rate 3.04×10^−3^ substitutions per site per year(95% BCI: 2·04×10^−3^ to 4·09×10^−3^ substitutions per site per year). Based on our demographic data, time interval from symptom onset to hospital admission was ∼8·5 days. The estimated reproduction number was 1.84(95% BCI: 1.37 to 2.35).

## Discussion

This study provides a territory-wide overview of early COVID-19 outbreak in Hong Kong, an international city with borders connecting to Mainland China, by integrating the demographic, clinical, epidemiological, phylogenomic, and phylodynamic data.

Fever was reported to be the most common symptom since 81·8%–98·0% of the patients had fever on admission during the initial outbreak in China(19-22). However, a large-scale study by Guan *et al* found that fever was only presented in 43·8% of the patients on admission(2). In this study, fever was identified in only 58·0% of the patients on admission, but developed in 64.0% after hospitalisation. Afebrile patients might be missed if the surveillance case definition relied solely on fever. Therefore, laboratory surveillance has been extended to in-patients and out-patients with respiratory symptoms. ICU admission was relatively uncommon(6·0%) in our cohort when compared with previous studies(19-21). Two ICU patients presented with severe complications including ARDS, acute renal damage, and septic shock. Of note, both patients had bacterial co-infection, which was absent in other cases. In addition to virulence factors of the pathogens, the host immune status, old age, and presence of chronic illness might be associated with enhanced disease severity. Immune supportive treatment and prompt antibiotics administration might reduce complications and mortality.

In Hong Kong, initial cases recorded in January 2020 were mostly imported cases. Since February 1, majority were local cases and their close contacts, indicating local community transmissions. Transmission in closed settings especially during family and religious gatherings is a hallmark of recent cases recorded in Hong Kong. Among six clusters identified based on epidemiological linkage, three(Clusters 4–6; figure 1) were considered SSEs with more individuals involved(n=8–13). WGS was performed on all 50 cases to investigate their phylogenetic relatedness and the transmission linkage.

The SARS-CoV-2 samples in Hong Kong have 99·98% identity to the reference genome(NC_045512.2), indicating that no major genome modification has occurred since the initial COVID-19 outbreak in Wuhan. Forty-four(88·0%) of our strains shared a common mutation *Orf3a* G251V. Except one imported case, all belonged to local cases and their close contacts without travel records(figure 1). Unlike other regions, such as China and Europe, where the SARS-CoV-2 genomes scattered across different branches in global phylogeny, the majority(88·0%) of Hong Kong strains were clustered in two subclades(figure 2). This suggested that the COVID-19 outbreak in Hong Kong community was mostly arisen from two ancestors.

Regarding the local phylogenetic analysis, clustering of samples was highly concordant to the epidemiological linkage. Cluster 1 demonstrated the closest genetic distance to the reference genome amongst all cases reported in Hong Kong(figures 1 and 3). The index case of Cluster 1(Case 66) was previously defined as possibly local infection as the patient travelled to Guangdong Province, which was not considered to have active community transmission by that time. However, our sequencing result showed that the genome of Case 66 was 100% identical to the Wuhan reference genome, and all cases in Cluster 1 did not harbour *Orf3a* G251V, which was recognized as a hallmark of the local cases with unknown source in our community. Therefore, instead of possibly local infections, Cluster 1 was more likely imported from Mainland China via index case 66.

Cluster 5 originated from a public estate, in which a family of three members(Cases 42, 48, and 49) was speculated to get infected from a confirmed case(Case 12) who lived in the same building, but 10-storey above, through a faulty sewage pipe setup. Based on phylogenetic analysis, viral genomes in Cluster 5 shared a similar genetic distance from the reference genome and were assigned to the same branch of the tree. This supports a potential transmission linkage among these cases.

Cluster 4 was a family gathering-associated SSE during Chinese New Year. In concert with epidemiological information, all 11 cases from Cluster 4 shared three common missense mutation, namely *S* gene L8V, *Orf1ab* H3233Y and *Orf3a* G251V, with seven sharing identical genomes. Considering the fast-evolving property of RNA viruses, identical genetic sequences among the strains implied that transmission occurred in a short period or even in a single event. Meanwhile, two singleton cases(Case 23 and Case 43) as well as cases from another local cluster(Case 38, Case 39, and Case 40) shared highly similar genomes to those of Cluster 4(figures 1 and 3). While no apparent epidemiological linkage were observed, the high degree of genome similarity suggests that these cases might be originated from a single source. The speculation was further supported by their geographical distribution of residences who lived in close proximity to each other, and might share overlapping living circles(figure 4).

**Figure 4.**
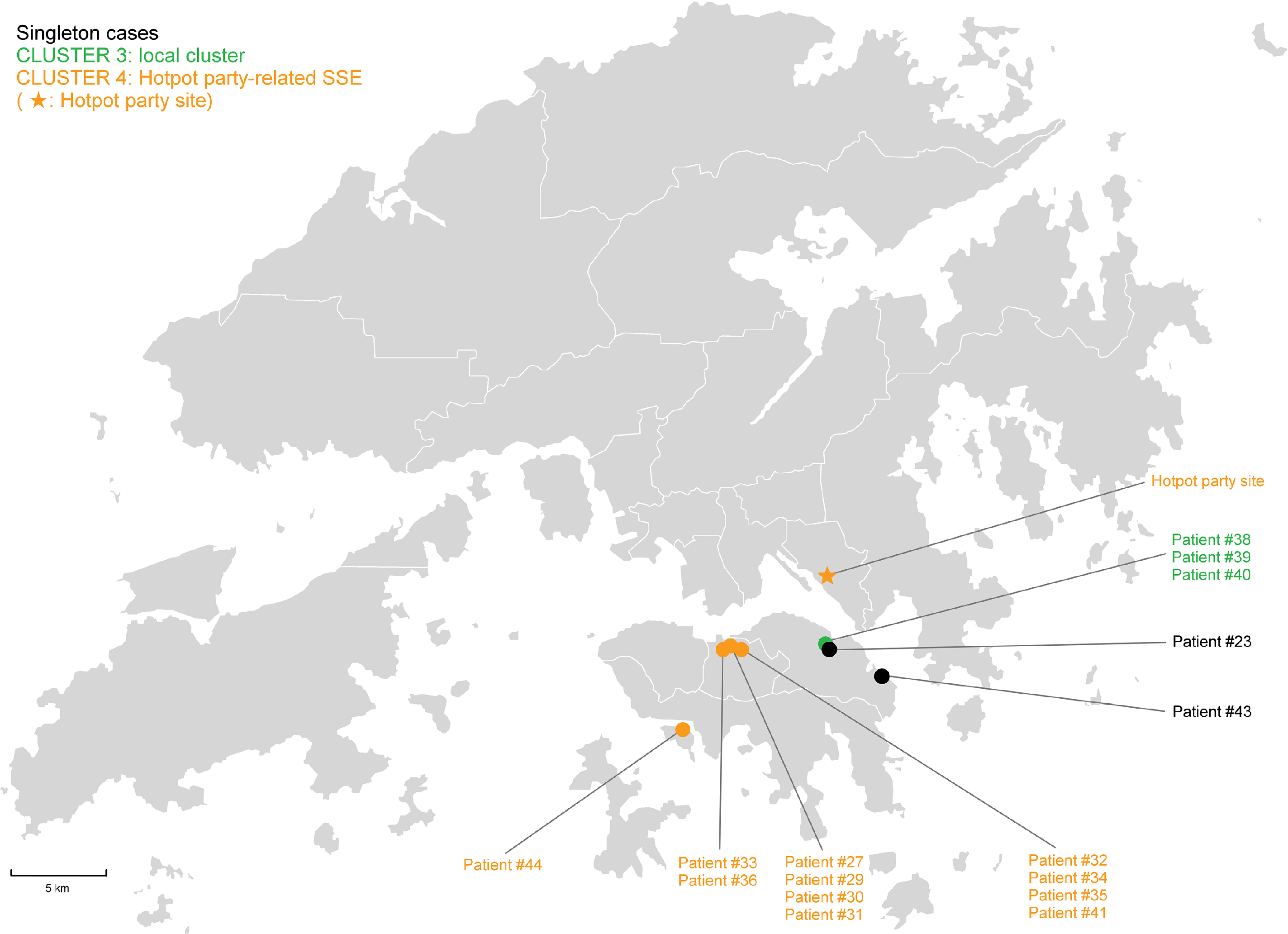
Geographic distribution of residences of singleton case #23, case #43, and cases in Cluster 3 and 4.

Cluster 6 was an SSE occurring in a Buddhist worship hall. Two missense mutations G295V and L3606F in *Orf1ab* were unique to this cluster. Epidemiological investigation identified a monk(Case 102; pink box of Cluster 6, figure 1), who was the abbot of the worship hall and had travelled to Mainland China in early January. He was sent to a quarantine centre in late February after being found associated with a series of confirmed cases connected with the worship hall. He was completely asymptomatic throughout the study period. Phylogenetic analysis showed that this case was closest to the root of the cluster(top portion with strains shown in red, figure 3), suggesting that Case 102 could be the index patient of the cluster. By the time of data cut-off, the cluster has already involved 13 patients and the spread was still ongoing. Here we demonstrated the possibility of “hidden spreader” as a source of COVID-19 community outbreak. It also highlights the importance of rapid quarantine of the close contacts of confirmed cases regardless of the presence of signs and symptoms in order to halt the COVID-19 community spread.

In the evolutionary clock study, death rate δ(which refers to the duration for the case to become non-contagious) was determined as the lag time between symptom onset date and the hospital admission date(i.e. 8·5 days, equivalent to 366/8.5=45·18/year). While δ was normally calculated based on recovery date(23), admission date was considered for calculation because the transmission link in Hong Kong was practically stopped once the patient was hospitalised. Assuming δ=45·18/year in our calculation, reproduction number within Hong Kong up to February 28, 2020 was estimated at 1.84(95% BCI: 1·37 to 2·35). The value strongly indicated that the outbreak in Hong Kong is still ongoing, but was smaller than the estimated reproduction number of 2·6 in Wuhan(24). The smaller value is a combined outcome of reduced growth rate and increased δ. The former one is attributed to very strong public health awareness among Hong Kong people with greatly reduced social activities and always put on surgical masks during this period,(25, 26) whereas the latter is the result of robust laboratory surveillance and fast quarantine time. In addition, tMRCA for the cases in Hong Kong was determined on December 24, 2019, which is ∼25 days since the first symptom onset case(Case 2–the case with the earliest symptom onset, January 18, 2020; figure 1) of our study cohort.

There are several limitations in our study. Despite 53·8% of the cases reported in Hong Kong up to February 28 were included, another 43 cases including two fatal cases were not analysed in this study. Moreover, incubation period of cases with unknown source might vary widely. Studies have shown that incubation period can vary from 4.5-15.8 days,(24) and can be even longer for patients presenting with mild symptoms. However, as the patient might already be infectious during incubation period, resultant reproductive number in this study could still be underestimated. Moreover, current calculation was solely based on phylodynamic analysis which could be different from those based on epidemiological models. Finally, gap regions were observed in some consensus genomes. This is mainly because WGS was performed on respiratory specimens instead of viral culture. The paucity of viral load in specimens could affect the yield of sequencing libraries. Nevertheless, the uncovered area only accounted for approximately 1-3% of the entire viral genome while the remaining mapped regions had an average coverage of >100X, which should provide sufficient and accurate information for our subsequent analyses.

In conclusion, phylogenomic data were consistent with the epidemiological findings that transmission in closed settings especially during family and religious gatherings is a hallmark of COVID-19 outbreak in Hong Kong. Social distancing and vigilant infection control measures, such as rapid isolation of suspected or confirmed cases and their close contacts, are crucial to the containment of COVID-19 transmission in the community.

## Data Availability

All the sequencing data (50 genomes of SARS-CoV-2) have been submitted to the GenBank with accession no. MT232662-MT232711.

## Acknowledgments

This study was supported by Faculty of Health and Social Science and Department of Health Technology and Informatics of The Hong Kong Polytechnic University. We appreciate Oxford Nanopore Technologies Limited, especially Hai WANG and Eva YU, for their supportive service in the delivery of sequencing flow cells and reagents, and the provision of technical advices.

## Author Bio

Dr. Kenneth Leung obtained his PhD in the Department of Microbiology, the University of Hong Kong in 2019. He received bioinformatic and phylogenetic analysis training under HKU Pasteur institute. His research focused on molecular diagnosis and epidemiology of emerging infectious disease such as *Mycobacterium tuberculosis* and HIV.

## Contributors

KSSL and GKHS designed the study, collected, analysed and interpreted the data, conducted literature search, and drafted and critically reviewed the manuscript; and GKHS was also responsible for securing funding for this study. TTLN, HYL, MPC, KKGT, and LKL conducted experiments and analysed the data. AKLW, MCYY, BKCW, AYMH, KTY, KCL, RWTL, EYKT, WSL, MCC, YYN, KMS, KSCF, SKYC, WKT, and TLQ were responsible for collecting and analysing the data, and finalizing the manuscript. DHKS and SPY analysed the data, and wrote and critically reviewed the manuscript. WCY were involved in data analysis, data interpretation and writing of the manuscript. All authors reviewed and approved the final version of the manuscript.

## Declaration of interests

We declare no competing interests.

## Notes

### Competing Interest Statement

The authors have declared no competing interest.

### Funding Statement

The project was supported by internal funds from the Faculty of Health and Social Science, and Department of Health Technology and Informatics of The Hong Kong Polytechnic University.

## References

1. Zhu N, Zhang D, Wang W, Li X, Yang B, Song J, et al. A Novel Coronavirus from Patients with Pneumonia in China, 2019. N Engl J Med. 2020 Feb 20;382(8):727–33.

2. Guan WJ, Ni ZY, Hu Y, Liang WH, Ou CQ, He JX, et al. Clinical Characteristics of Coronavirus Disease 2019 in China. N Engl J Med. 2020 Feb 28.

3. Chan JF, Yuan S, Kok KH, To KK, Chu H, Yang J, et al. A familial cluster of pneumonia associated with the 2019 novel coronavirus indicating person-to-person transmission: a study of a family cluster. Lancet. 2020 Feb 15;395(10223):514–23.

4. Li Q, Guan X, Wu P, Wang X, Zhou L, Tong Y, et al. Early Transmission Dynamics in Wuhan, China, of Novel Coronavirus-Infected Pneumonia. N Engl J Med. 2020 Jan 29.

5. Dong E, Du H, Gardner L. An interactive web-based dashboard to track COVID-19 in real time. Lancet Infect Dis. 2020 Feb 19.

6. Centre_for_Health_Protection. Latest local situation of Severe Respiratory Disease associated with a Novel Infectious Agent. 2020 [cited 2020 24 February]; Available from: https://www.chp.gov.hk/files/pdf/enhanced_sur_pneumonia_wuhan_eng.pdf

7. Leung YF, Wong HB, Shing Y, Wong HW, Wong WK. wars.vote4.hk – Coronavirus in HK. 2020 [cited 2020 25 February]; Available from: https://wars.vote4.hk/en/

8. Centre_for_Health_Protection. Severe respiratory disease associated with a novel infectious agent—letters to doctors. 2020 [cited 2020 6 February]; Available from: https://www.chinadaily.com.cn/a/202001/25/WS5e2bb1b6a31012821727333a.html

9. Corman VM, Landt O, Kaiser M, Molenkamp R, Meijer A, Chu DK, et al. Detection of 2019 novel coronavirus (2019-nCoV) by real-time RT-PCR. Euro Surveill. 2020 Jan;25(3).

10. WHO. Clinical management of severe acute respiratory infection when Novel coronavirus (nCoV) infection is suspected: interim guidance. 2020 [cited 2020 11 Jan]; Available from: https://www.who.int/internal-publications-detail/clinical-management-of-severe-acute-respiratory-infection-when-novel-coronavirus-(ncov)-infection-is-suspected

11. Peiris JS, Lai ST, Poon LL, Guan Y, Yam LY, Lim W, et al. Coronavirus as a possible cause of severe acute respiratory syndrome. Lancet. 2003 Apr 19;361(9366):1319–25.

12. Quick J. Artic Network-nCoV 2019 sequencing protocol. 2020 [cited 2020 24 February]; Available from: https://artic.network/ncov-2019

13. Loman N, Rambaut A. nCoV-2019 novel coronavirus bioinformatics protocol. ARTIC-nCoV-bioinformaticsSOP-v100 2020 [cited 2020 10 March]; Available from: https://artic.network/ncov-2019/ncov2019-bioinformatics-sop.html

14. DePristo MA, Banks E, Poplin R, Garimella KV, Maguire JR, Hartl C, et al. A framework for variation discovery and genotyping using next-generation DNA sequencing data. Nat Genet. 2011 May;43(5):491–8.

15. Guindon S, Dufayard JF, Lefort V, Anisimova M, Hordijk W, Gascuel O. New algorithms and methods to estimate maximum-likelihood phylogenies: assessing the performance of PhyML 3.0. Syst Biol. 2010 May;59(3):307–21.

16. Elbe S, Buckland-Merrett G. Data, disease and diplomacy: GISAID’s innovative contribution to global health. Glob Chall. 2017 Jan;1(1):33–46.

17. Bouckaert R, Heled J, Kuhnert D, Vaughan T, Wu CH, Xie D, et al. BEAST 2: a software platform for Bayesian evolutionary analysis. PLoS Comput Biol. 2014 Apr;10(4):e1003537.

18. Rambaut A, Drummond AJ, Xie D, Baele G, Suchard MA. Posterior Summarization in Bayesian Phylogenetics Using Tracer 1.7. Syst Biol. 2018 Sep 1;67(5):901–4.

19. Chen N, Zhou M, Dong X, Qu J, Gong F, Han Y, et al. Epidemiological and clinical characteristics of 99 cases of 2019 novel coronavirus pneumonia in Wuhan, China: a descriptive study. Lancet. 2020 Feb 15;395(10223):507–13.

20. Huang C, Wang Y, Li X, Ren L, Zhao J, Hu Y, et al. Clinical features of patients infected with 2019 novel coronavirus in Wuhan, China. Lancet. 2020 Feb 15;395(10223):497–506.

21. Wang D, Hu B, Hu C, Zhu F, Liu X, Zhang J, et al. Clinical Characteristics of 138 Hospitalized Patients With 2019 Novel Coronavirus-Infected Pneumonia in Wuhan, China. JAMA. 2020 Feb 7.

22. Wu J, Liu J, Zhao X, Liu C, Wang W, Wang D, et al. Clinical Characteristics of Imported Cases of COVID-19 in Jiangsu Province: A Multicenter Descriptive Study. Clin Infect Dis. 2020 Feb 29.

23. Boskova V, Stadler T, Magnus C. The influence of phylodynamic model specifications on parameter estimates of the Zika virus epidemic. Virus Evol. 2018 Jan;4(1):vex044.

24. Lai A, Bergna A, Acciarri C, Galli M, Zehender G. Early phylogenetic estimate of the effective reproduction number of SARS-CoV-2. J Med Virol. 2020 Feb 25.

25. Sung AD, Sung JAM, Thomas S, Hyslop T, Gasparetto C, Long G, et al. Universal Mask Usage for Reduction of Respiratory Viral Infections After Stem Cell Transplant: A Prospective Trial. Clin Infect Dis. 2016 Oct 15;63(8):999–1006.

26. Seto WH, Tsang D, Yung RW, Ching TY, Ng TK, Ho M, et al. Effectiveness of precautions against droplets and contact in prevention of nosocomial transmission of severe acute respiratory syndrome (SARS). Lancet. 2003 May 3;361(9368):1519–20.

